# Global and regional burden of attributable and associated bacterial antimicrobial resistance avertable by vaccination: modelling study

**DOI:** 10.1101/2022.05.08.22274821

**Authors:** Chaelin Kim, Marianne Holm, Isabel Frost, Mateusz Hasso-Agopsowicz, Kaja Abbas

**Author notes:** **Corresponding author:** Chaelin Kim ( /).

## Abstract

**Introduction:** Antimicrobial resistance (AMR) is a global health threat with 1.27 million and 4.95 million deaths attributable to and associated with bacterial AMR respectively in 2019. Our aim is to estimate the vaccine avertable bacterial AMR burden based on existing and future vaccines at the regional and global levels by pathogen and infectious syndromes.

**Methods:** We developed a static proportional impact model to estimate the vaccination impact on 15 bacterial pathogens in terms of reduction in age-specific AMR burden estimates for 2019 from the Global Research on Antimicrobial Resistance project in direct proportion to efficacy, coverage, target population for protection, and duration of protection of existing and future vaccines.

**Results:** In the baseline scenario for vaccination of primary age-groups against 15 pathogens, we estimated vaccine-avertable AMR burden of 0.51 (95% UI: 0.49 - 0.54) million deaths and 28 (27 - 29)million DALYs associated with bacterial AMR, and 0.15 (0.14 - 0.17) million deaths and 7.6 (7.1 - 8.0) million DALYs attributable to AMR globally in 2019. In the high-potential scenario for vaccination of additional age groups against 7 pathogens, we estimated vaccine-avertable AMR burden of an additional 1.2 (1.18 - 1.23) million deaths and 37 (36 - 39) million DALYs associated with AMR, and 0.33 (0.32 - 0.34) million deaths and 10 (9.8 - 11) million DALYs attributable to AMR globally in 2019.

**Conclusion:** The AMR burden avertable by vaccination in 2019 was highest for the WHO Africa and South-East Asia regions, for lower respiratory infections, tuberculosis, and bloodstream infections by infectious syndromes, and for *Mycobacterium tuberculosis* and *Streptococcus pneumoniae* by pathogen. Increased coverage of existing vaccines and development of new vaccines are effective means to reduce AMR, and this evidence should inform the full value of vaccine assessments.

**Key questions:** *What is already known on this topic:*

- There is some evidence on the impact of vaccines against *Haemophilus influenzae* type b, rotavirus, *Streptococcus pneumoniae, Salmonella* Typhi and influenza on antimicrobial resistance (AMR) in specific settings.

*What this study adds:*

- To our knowledge, this is the first study to estimate attributable and associated bacterial AMR burden avertable by vaccination against 15 bacterial pathogens for a combined set of existing and new vaccines in the pipeline by pathogen, infectious syndrome, and region.
- The AMR burden avertable by vaccination in 2019 was highest for the WHO Africa and South-East Asia regions, for lower respiratory infections, tuberculosis, and bloodstream infections by infectious syndromes, and for *Mycobacterium tuberculosis* and *Streptococcus pneumoniae* by pathogen.

*How this study might affect research, practice or policy:*

- Our model-based projections facilitate evidence-based decision-making for scaling up of existing vaccines to regions in most need with higher AMR burden and prioritise development of new vaccines with high potential for lowering AMR burden by pathogen, infectious syndrome, and region.
- Our study contributes to the WHO-led value attribution framework for vaccines against antimicrobial resistance, and specifically to the criterion focused on vaccine averted AMR health burden.

## Introduction

Since the discovery of penicillin in 1928, antimicrobials have been used to treat bacteria, fungi, parasites, and viruses, saving countless lives [1]. However, antimicrobial resistance (AMR) is a growing global public health threat in the 21st century [2]. Resistance occurs through pathogen evolution, either naturally over time or acquired by the use of antimicrobial drugs, which render these drugs ineffective and increase the risk of morbidity and mortality. While access to antimicrobial drugs in low- and middle-income countries to treat infections continues to be a challenge, misuse and overuse of antimicrobials along with lack of access to clean water, sanitation and hygiene (WASH) and effective infection prevention and control (IPC) measures have fuelled the emergence and spread of AMR globally. The UK government commissioned review on AMR in 2014 projected that if AMR is not controlled, it would lead to significant impact on health with 10 million AMR-related deaths annually and macroeconomic consequences with a cumulative economic loss of US$ 100 trillion by 2050 [3].

Vaccination, when used in conjunction with other preventive measures, has the potential to significantly reduce AMR transmission through several pathways [4,5]. First, vaccination has a direct influence on the health burden of AMR by preventing the emergence and transmission of drug-resistant and drug-sensitive infections, and the associated antibiotic use. Second, vaccines have an indirect influence by reducing resistant infections in unvaccinated populations through herd immunity. Third, vaccination can prevent infections where antimicrobials are not indicated but often wrongly prescribed, such as primary viral infections, thereby reducing misuse and overuse of antimicrobials. Fourth, vaccines can also reduce the use of antimicrobials to treat secondary bacterial infections caused by viral diseases. Finally, vaccines can give longer-term health benefits in preventing infections and resistance to vaccines is rarely observed [6].

The Global Research on Antimicrobial Resistance (GRAM) project estimated the deaths and disability-adjusted life-years (DALYs) attributable to and associated with resistance by replacing all drug-resistant infections with susceptible infection or no infection, respectively. It estimated that 1.27 (95% UI: 0.91 - 1.7) million deaths and 47.9 (35 - 64) million DALYs were attributable to bacterial AMR and 4.95 (3.6 - 6.6) million deaths and 192 (146 - 248) million DALYs were associated with bacterial AMR in 2019 [7]. Despite the significant potential impact of vaccination in lowering AMR, evidence is limited due to the methodological difficulties and challenges in obtaining data on the health burden associated with AMR in order to calculate this impact [8–10]. Such evidence will be valuable to inform improvements in the coverage of existing vaccines and prioritise research and development of new vaccines.

To address this evidence gap, our aim is to analyse the findings from the GRAM project and estimate the vaccine-avertable bacterial AMR burden based on the profiles of existing and future vaccines by pathogen and infectious syndromes at the regional and global levels in 2019. Such pan-pathogen analyses using standardised approaches are critical to inform vaccine development, funding, introduction and use. They also inform the WHO-led value attribution framework for vaccines against AMR [11], which includes five criteria: (i) vaccine averted AMR health burden, (ii) vaccine averted AMR economic burden, (iii) vaccine averted antibiotic use, (iv) sense of urgency to develop antimicrobial approaches, and (v) pathogen impact on equity and social justice. Our study contributes to the first criterion – vaccine-averted AMR health burden.

## Methods

### AMR burden data

We used the bacterial AMR burden estimates from the Global Research on Antimicrobial Resistance (GRAM) project which provided data for age-specific deaths and DALYs associated with and attributable to AMR by pathogen, infectious syndrome, and region for 2019 [7]. These comprehensive estimates of bacterial AMR burden were based on statistical predictive modelling of data from systematic reviews, surveillance systems, hospital systems, and other sources to generate estimates for 23 pathogens and 88 pathogen-drug combinations for 204 countries in 2019. The AMR burden estimates for *Neisseria gonorrhoeae* includes only morbidity and no mortality.

Two sets of estimates are presented – burden attributed to AMR, that is deaths and DALYs that could be averted if all drug-resistant infections would be replaced by drug-sensitive infections; and burden associated with AMR, that is deaths and DALYs that could be averted if all drug-resistant infections would be replaced by no infections. As vaccines prevent drug-resistant and drug-susceptible burden, we infer that the associated AMR burden is the appropriate metric for measuring the impact of vaccination on AMR burden.

### Vaccine profiles

We focused our analysis on 15 pathogens – *Acinetobacter baumannii, Enterococcus faecium, Escherichia coli*, Group A *Streptococcus, Haemophilus influenzae, Klebsiella pneumoniae, Mycobacterium tuberculosi*s, *Neisseria gonorrhoeae*, non-typhoidal *Salmonella, Pseudomonas aeruginosa, Salmonella* Paratyphi, *Salmonella* Typhi, *Shigella* spp., *Staphylococcus aureus*, and *Streptococcus pneumoniae*. We selected pathogens that are part of the WHO evaluation of the value of vaccines in preventing AMR. We used vaccine profiles (see Table 1), which comprise the vaccine target population, efficacy, coverage, duration of protection, and disease presentation prevented.

**Table 1.**
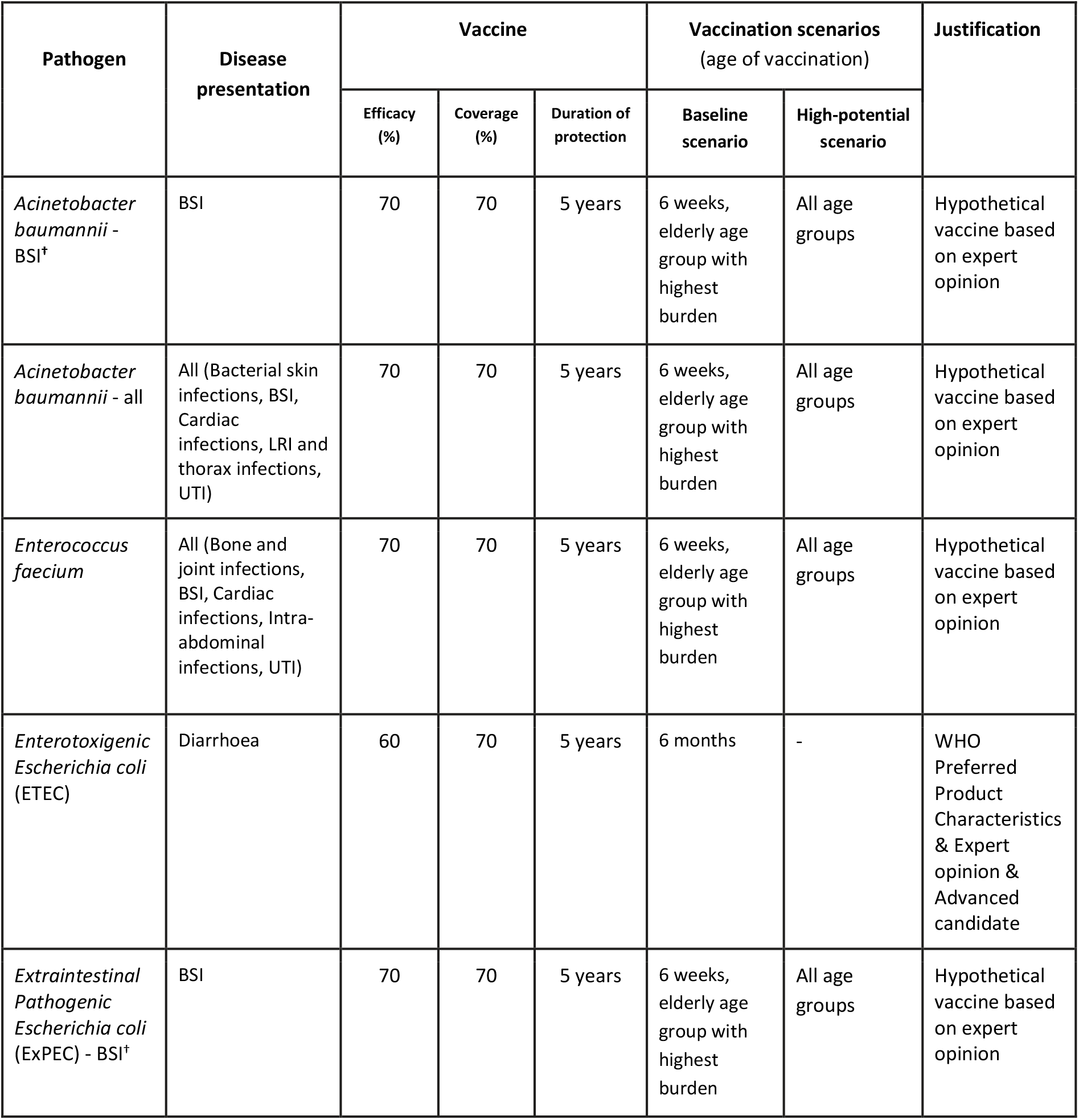
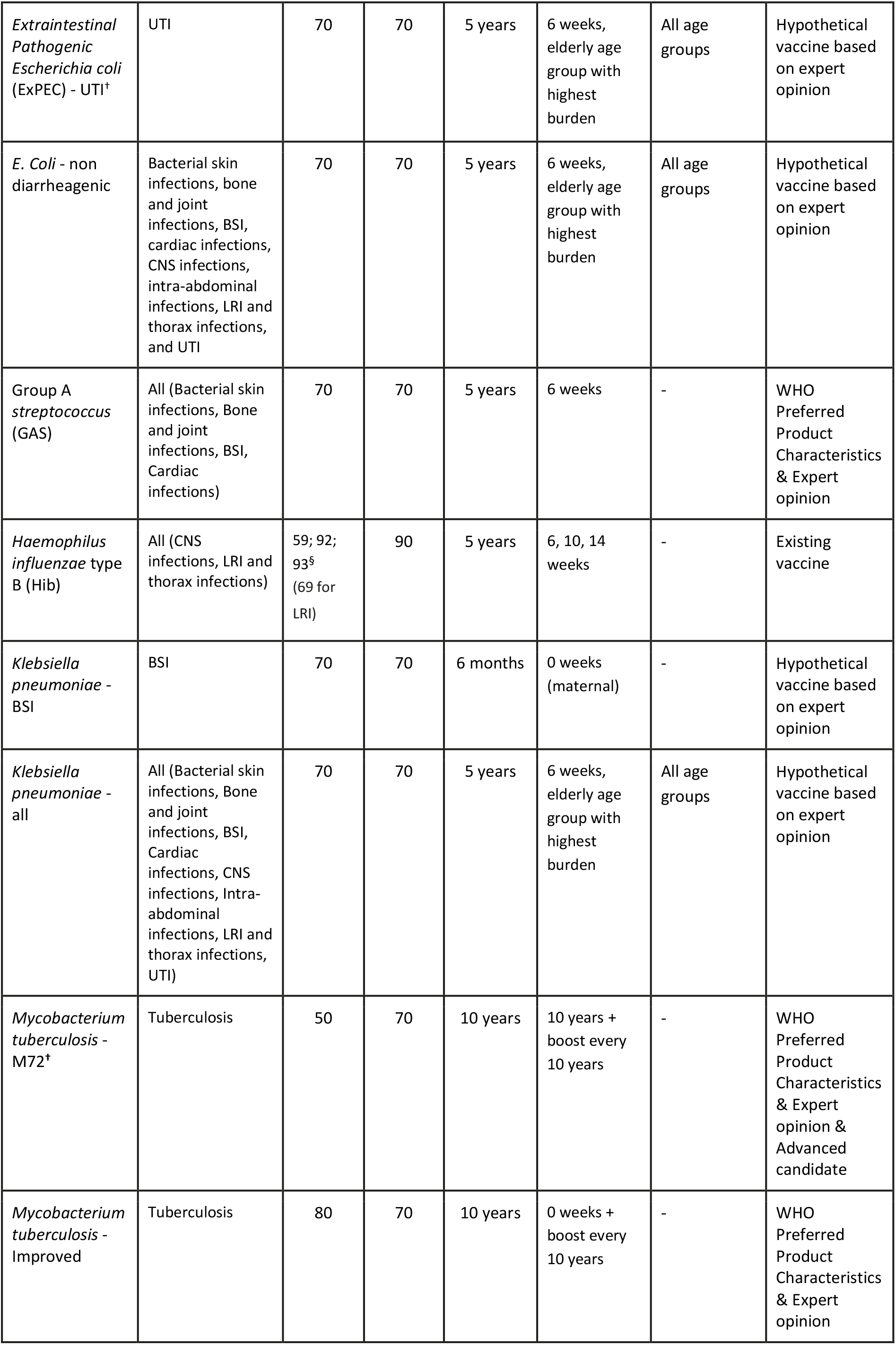
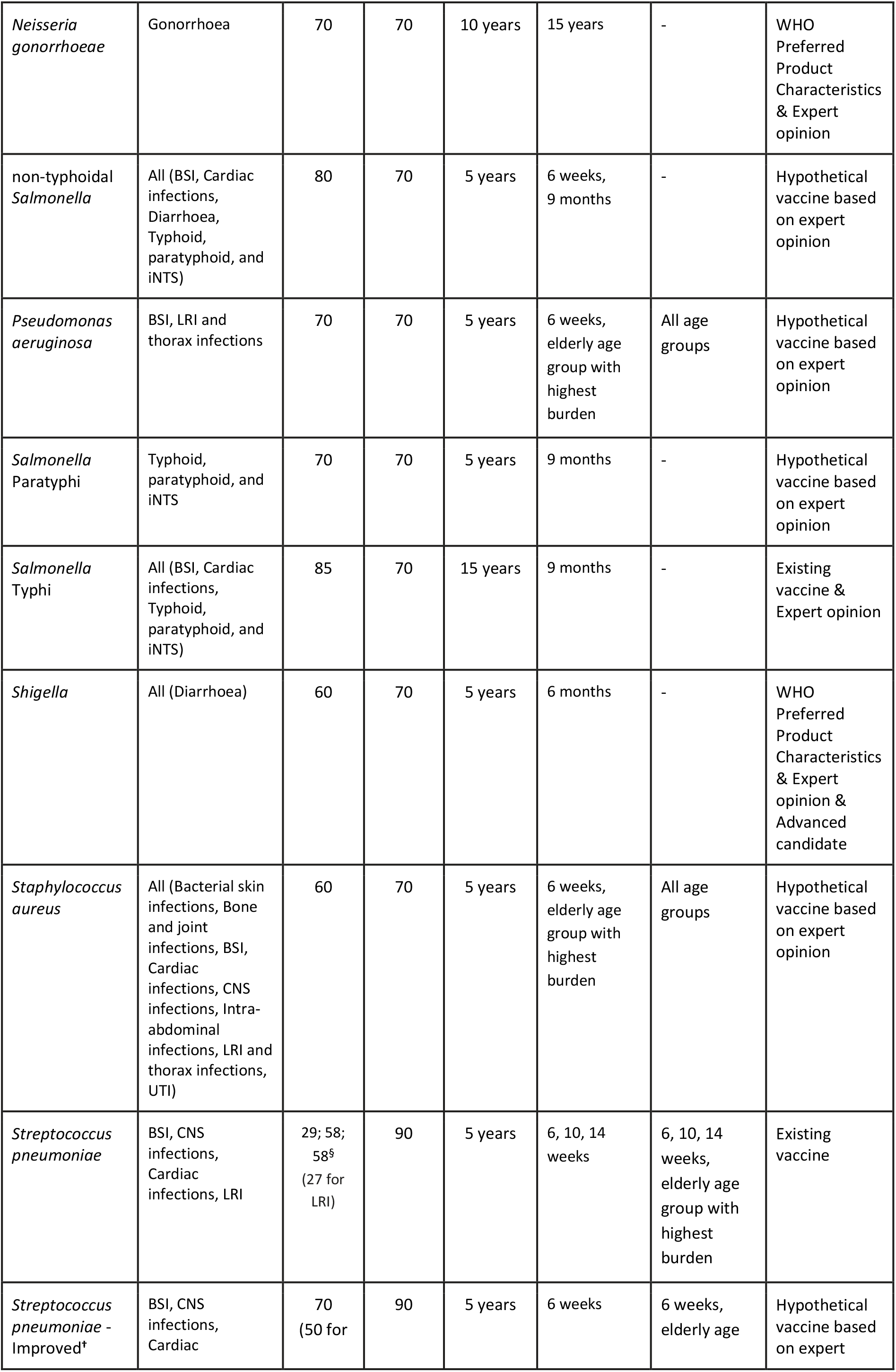
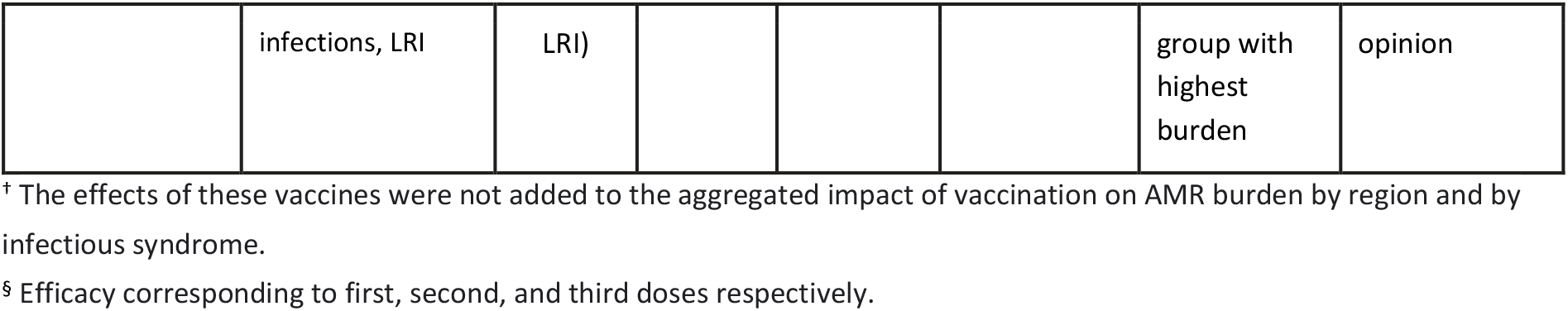
Vaccine profiles. Product characteristics for efficacy and duration of protection for vaccine-derived immunity, and coverage and target population (age of vaccination) for current and future vaccines against bacterial pathogens. The baseline scenario includes 15 pathogens, and the high-potential scenario includes a subset of 7 pathogens. (Bone and joint infections = infections of bones, joints, and related organs; BSI = bloodstream infections; cardiac infections = endocarditis and other cardiac infections; CNS infections = meningitis and other bacterial CNS infections; intra-abdominal infections = peritoneal and intra-abdominal infections; LRI and thorax infections = lower respiratory infections and all related infections in the thorax; bacterial skin infections = bacterial infections of the skin and subcutaneous systems; typhoid, paratyphoid, and iNTS = typhoid fever, paratyphoid fever, and invasive non-typhoidal *Salmonella* spp; UTI = urinary tract infections and pyelonephritis)

For the existing vaccines against *H. influenzae* type b, *Streptococcus pneumoniae*, and *Salmonella* Typhi, the vaccine profiles expand coverage of the current vaccines in order to meet the strategic priority on coverage and equity of Immunisation Agenda 2030 [12]. For vaccines that are not yet available, hypothetical profiles were developed based on preferred product characteristics (PPCs) [13], target product profiles (TPPs), attributes of advanced vaccine candidates, and expert consultations with WHO working groups, PATH, and pathogen experts. Some pathogens have multiple disease presentations and would require different vaccines to prevent different disease presentations. As such, these pathogens have more than one vaccine profile.

### Modelling process

We developed a static proportional impact model to estimate the vaccination impact in terms of reduction in age-specific AMR burden estimates for 2019 from the GRAM project. We estimated a counterfactual pre-vaccination scenario for diseases with current vaccinations and adjusted for disease type specification before applying the vaccine impact. We calculated the reduction in pre-vaccine AMR burden after vaccination in direct proportion to efficacy, coverage, target population for protection, and duration of protection of existing and potential future vaccines [14].

For ages that lie within the duration of protection since the time of vaccination:

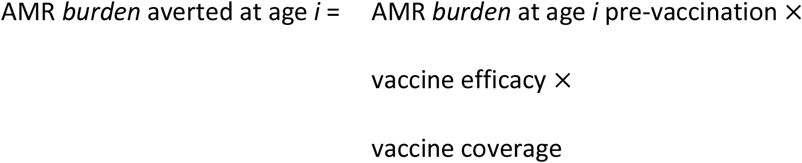

### Scenarios

We estimated vaccine avertable deaths and DALYs attributable to and associated with AMR by region, infectious syndrome, and pathogen with 95% uncertainty intervals (UIs) for two scenarios – baseline scenario (for 15 pathogens) for primary vaccination of specific age-groups, and high-potential scenario (for a subset of 7 pathogens) that includes additional age groups at risk of infection based on expert opinion.

Vaccine profiles with the corresponding product characteristics for efficacy and duration of protection for vaccine-derived immunity, and coverage and target population for the baseline and high-potential scenarios are described in Table 1. In the *baseline* scenario, we estimated the vaccine avertable burden from the age of vaccination under the assumption that vaccine-derived immunity would sustain for the duration of protection of the corresponding vaccines. We did not consider vaccine waning dynamics due to limited evidence. For pathogens with a highly uncertain vaccine target population or feasibility of vaccine delivery, we estimated an additional *high-potential* scenario which assumed that individuals at risk (including additional age groups at risk) would be vaccinated to protect against corresponding disease presentations. This was applicable to vaccines against *Acinetobacter baumannii, Enterococcus faecium*, Extraintestinal Pathogenic *Escherichia coli* (ExPEC), *Klebsiella pneumoniae (all syndromes), Pseudomonas aeruginosa*, and *Staphylococcus aureus*. For *Streptococcus pneumoniae*, we explored the high-potential scenario by administering a vaccine to an elderly population with the highest disease burden.

### Uncertainty analysis

We conducted a Monte Carlo simulation of 400 runs (sufficient for results to converge) to propagate the uncertainty in the AMR burden, vaccine efficacy, and coverage through the model simulations to estimate the uncertainty in our projected outcomes of vaccination impact. We provide summary estimates in terms of vaccine-avertable deaths and DALYs attributable to and associated with AMR by region, infectious syndrome, and pathogen with 95% uncertainty intervals (UIs) for the baseline and high-potential scenarios. Additional details on the modelling process, scenarios, and uncertainty analysis are provided in the Appendix A1.

## Results

### Vaccine impact on global AMR burden

At the global level in 2019 for the baseline scenario, we estimated that vaccines against the 15 pathogens (analysed in this study) could avert 0.51 (95% UI: 0.49 - 0.54) million deaths and 28 (27 - 29) million DALYs associated with AMR, and 0.15 (0.14 - 0.17) million deaths and 7.6 (7.1 - 8.0) million DALYs attributable to AMR. In the high-potential scenario, we estimated that vaccines against a subset of 7 pathogens could avert an additional 1.2 (1.18 - 1.23) million deaths and 37 (36 - 39) million DALYs associated with AMR, and 0.33 (0.32 - 0.34) million deaths and 10 (9.8 - 11) million DALYs attributable to AMR globally in 2019.

### Vaccine impact on AMR burden by pathogen

Figure 1a and Table 2a present the vaccine avertable burden attributable to and associated with AMR in 2019 for each of the pathogen-specific vaccine profiles at the global level for the baseline scenario. For pathogens with licensed vaccines, we estimated that vaccination against *Streptococcus pneumoniae* at 2019 coverage levels averted 44 (37 - 52) thousand deaths and 3.8 (3.3 - 4.5) million DALYs associated with AMR in 2019. By reaching the WHO recommended coverage level of 90% globally, 59 (50 - 69) thousand deaths and 5.1 (4.5 - 5.9) million DALYs associated with AMR could have been averted in 2019. Expanding the coverage to elderly populations would increase the vaccination impact to avert 71 (63 - 81) thousand deaths. We estimated that vaccination against *H. influenzae* at 2019 coverage levels averted 11 (9.7 - 13) thousand deaths and 0.98 (0.85 - 1.2) million DALYs associated with AMR in 2019. At 90% coverage globally, 13 (11 - 15) thousand deaths and 1.1 (0.96 - 1.3) million DALYs associated with AMR could have been averted. We estimated that wider introduction and scale-up of vaccination against *Salmonella* Typhi could have averted 34 (26 - 44) thousand deaths and 2.8 (2.2 - 3.6) million DALYs associated with AMR in 2019.

**Figure 1.**
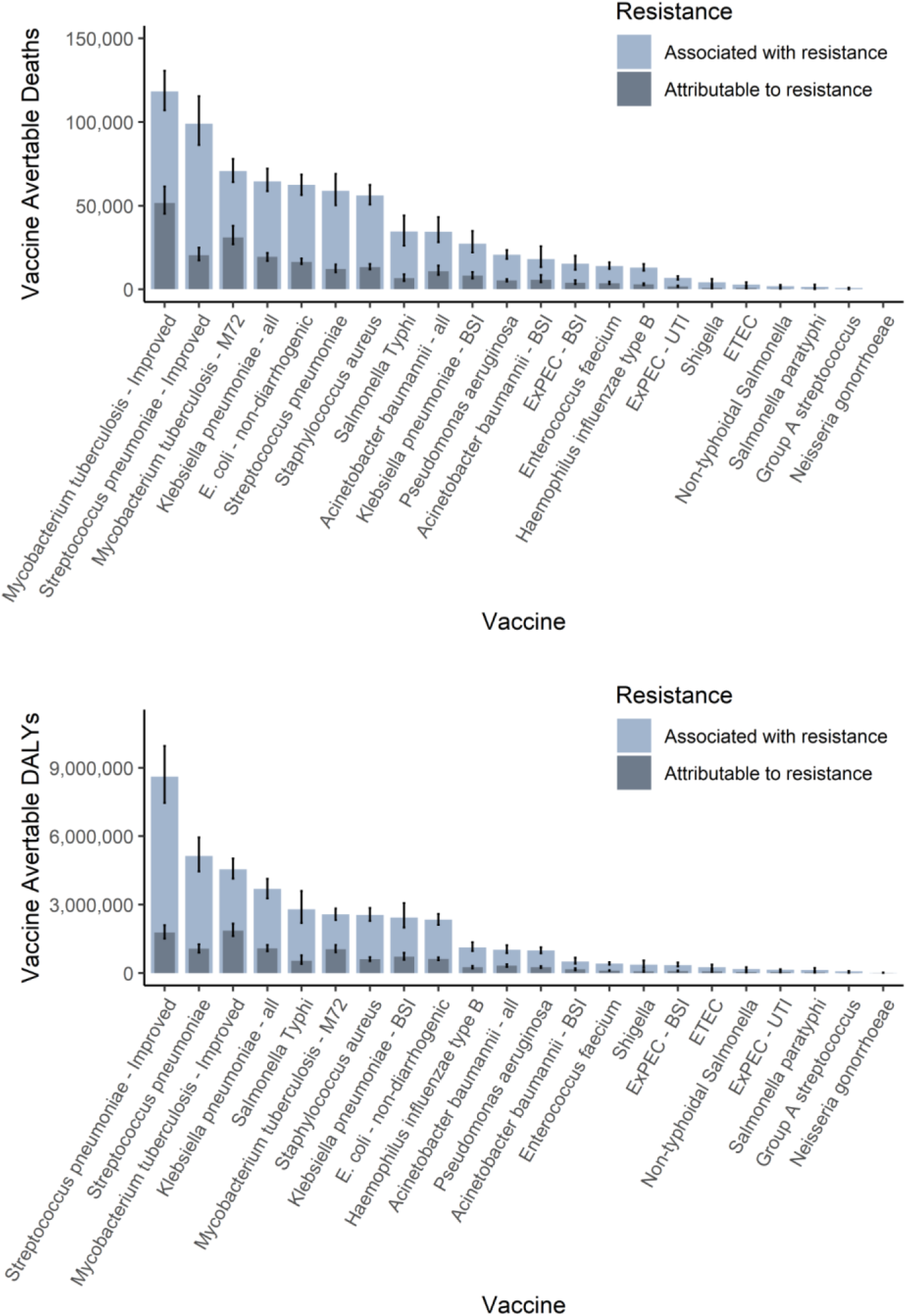
Vaccine impact on AMR burden by pathogen, infectious syndrome, and region. **Figure 1a. Vaccine impact on AMR burden by pathogen-specific vaccine profile**. The estimates (median and 95% uncertainty intervals) of the vaccine avertable deaths attributable to and associated with bacterial antimicrobial resistance in 2019 were aggregated by vaccine profile in the baseline scenario.

**Figure 1b.**
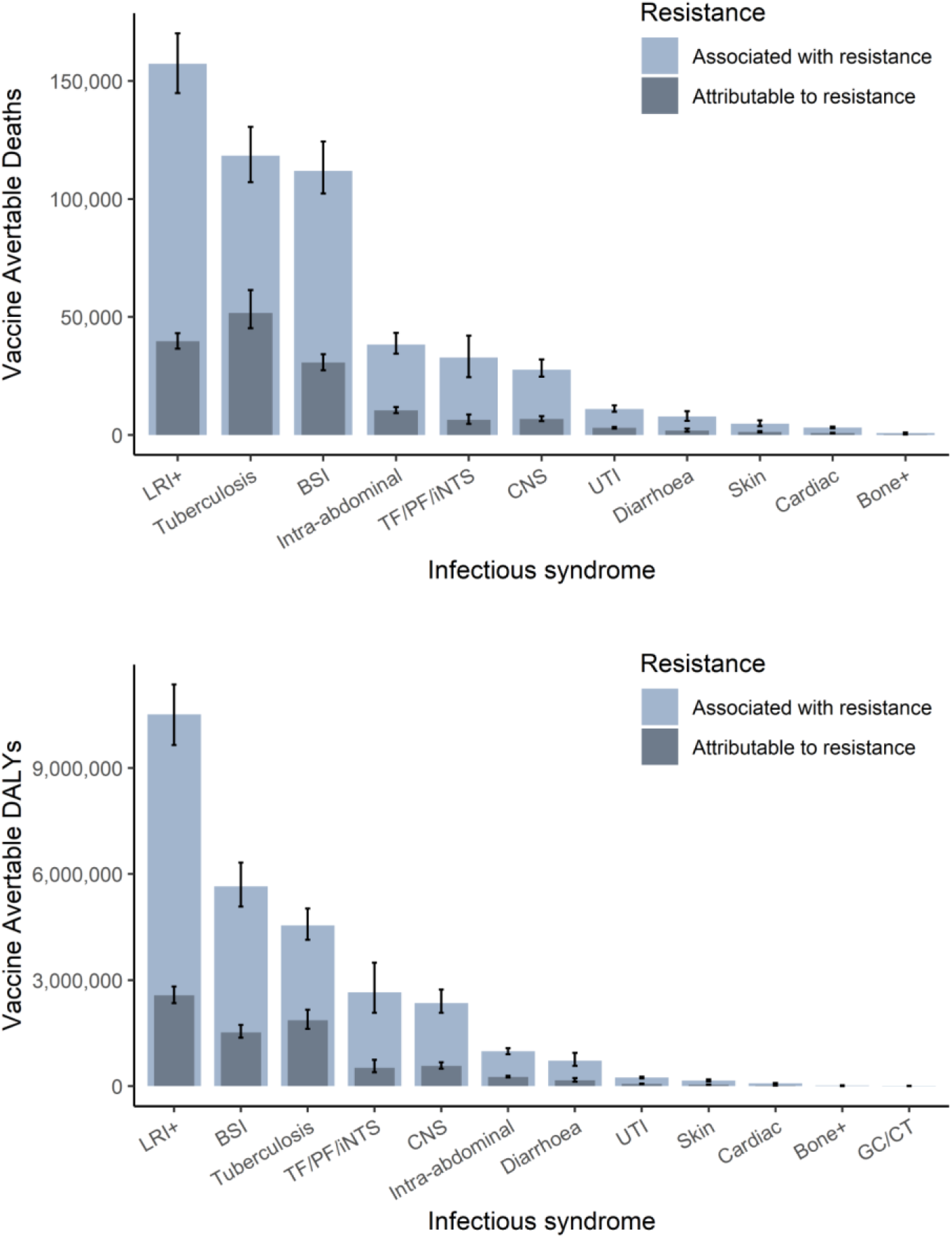
Vaccine impact on AMR burden by infectious syndrome. The estimates (median and 95% uncertainty intervals) of the vaccine avertable deaths attributable to and associated with bacterial antimicrobial resistance in 2019 were aggregated by infectious syndrome in the baseline scenario. (Bone+ = infections of bones, joints, and related organs; BSI = bloodstream infections; cardiac = endocarditis and other cardiac infections; CNS = meningitis and other bacterial CNS infections; intra-abdominal = peritoneal and intra-abdominal infections; LRI+ = lower respiratory infections and all related infections in the thorax; skin = bacterial infections of the skin and subcutaneous systems; TF–PF–iNTS = typhoid fever, paratyphoid fever, and invasive non-typhoidal *Salmonella* spp; UTI = urinary tract infections and pyelonephritis)

**Figure 1c.**
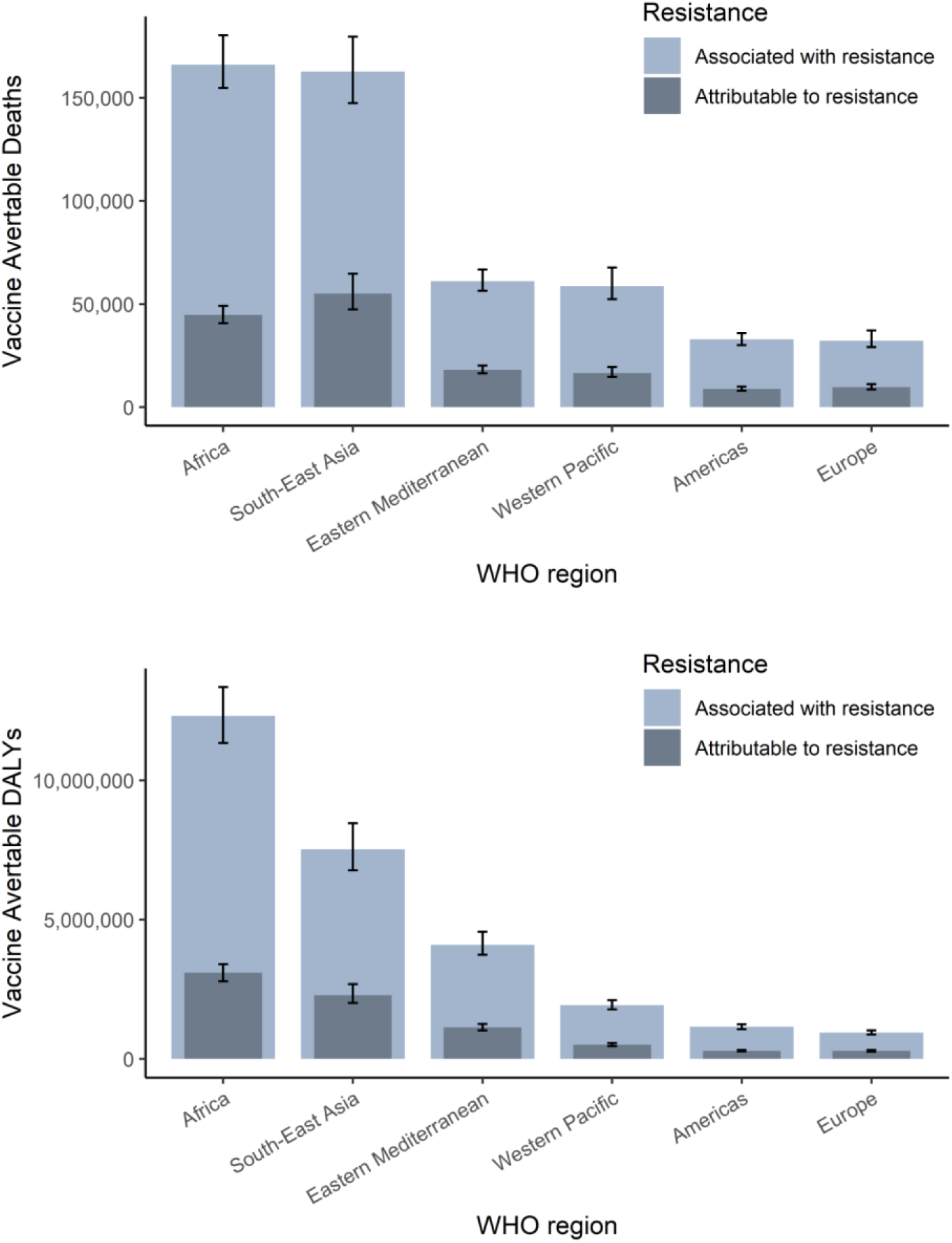
Vaccine impact on AMR burden by WHO region. The estimates (median and 95% uncertainty intervals) of the vaccine avertable deaths attributable to and associated with bacterial antimicrobial resistance in 2019 were aggregated by WHO region in the baseline scenario.

**Table 2.**
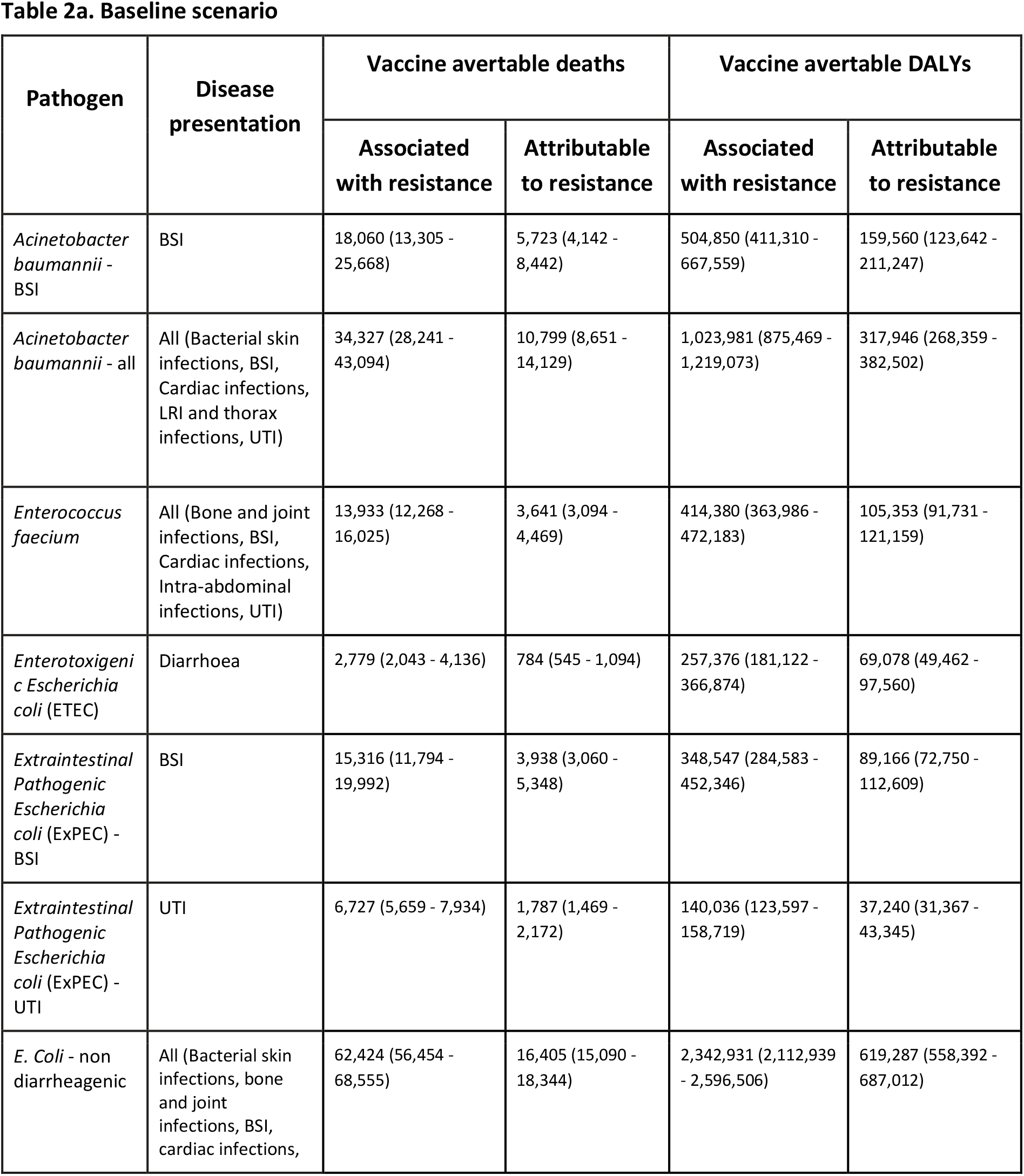
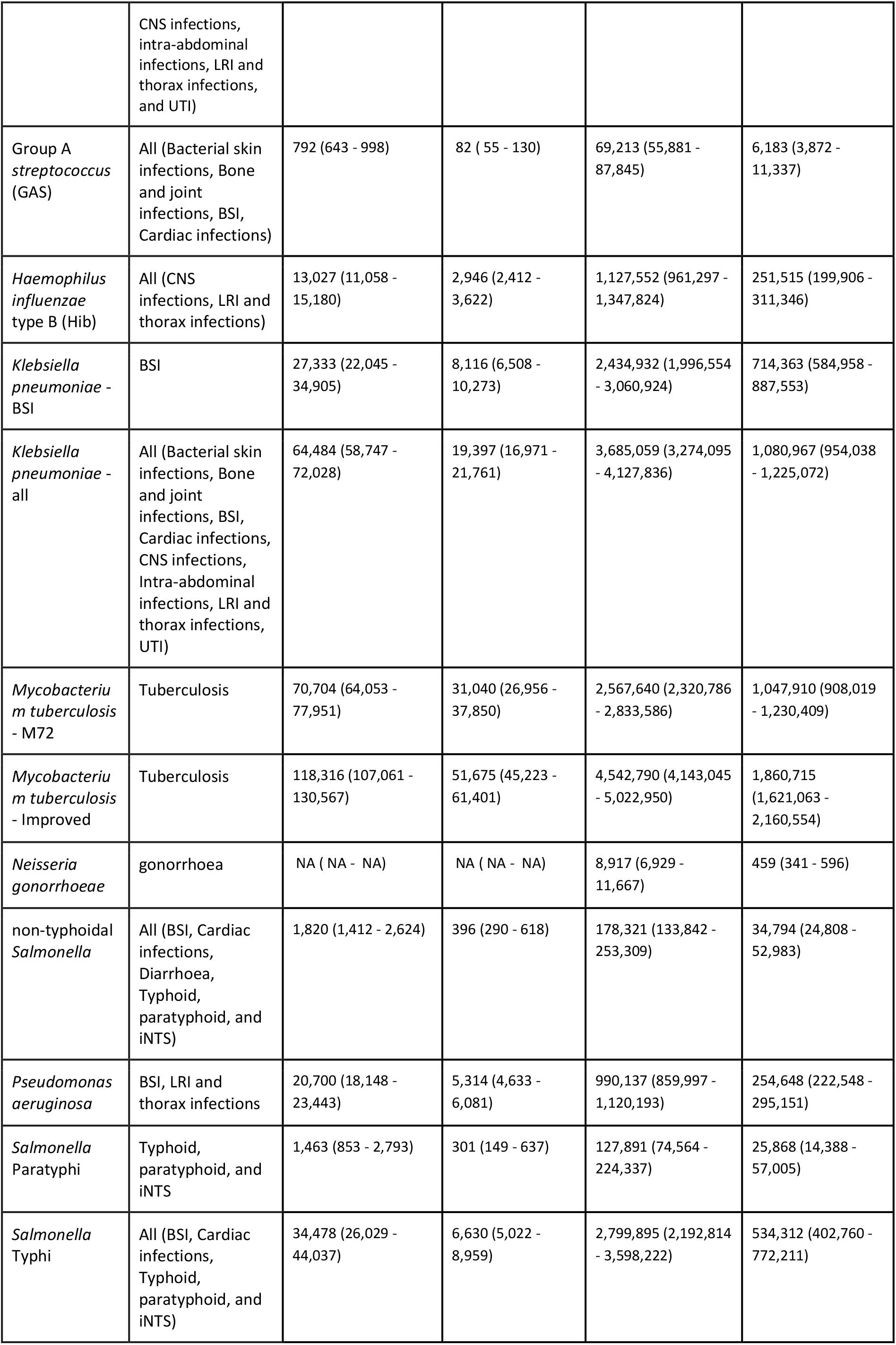
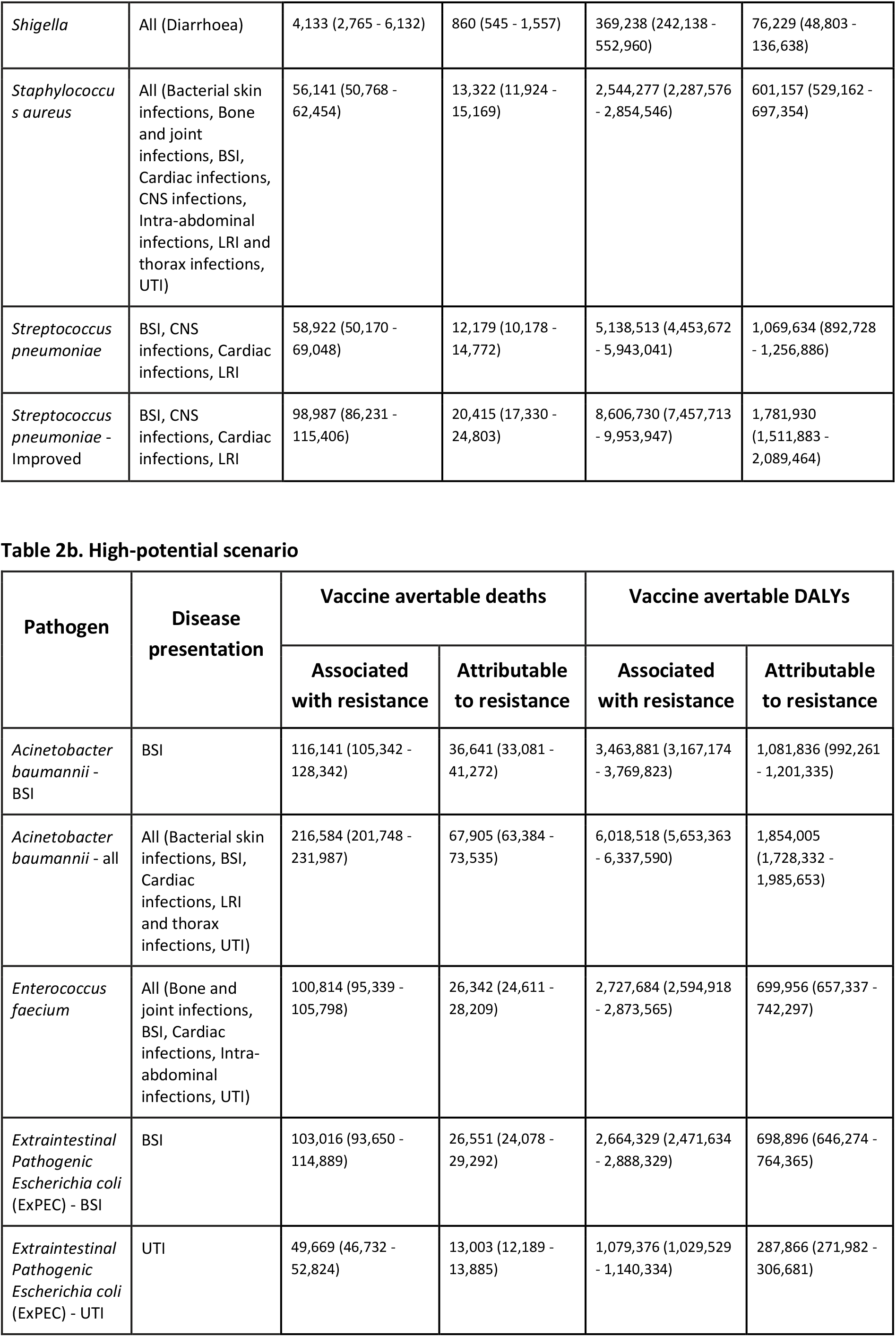
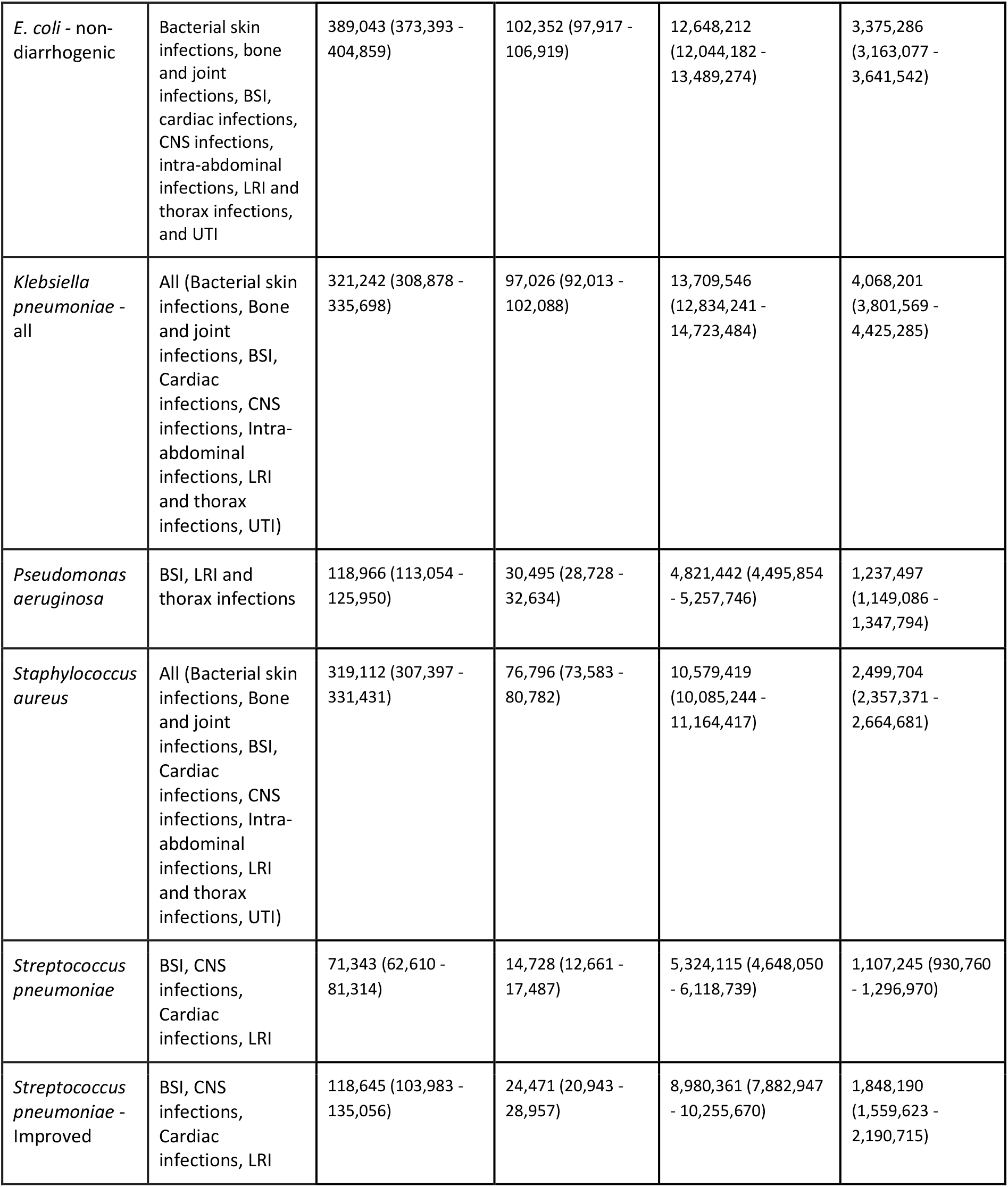
Vaccine impact on AMR burden by vaccine profile. The estimates (median and 95% uncertainty intervals) of the vaccine avertable deaths and DALYs attributable to and associated with bacterial antimicrobial resistance in 2019 were aggregated by vaccine profile for the baseline (Table 2a) and high-potential (Table 2b) scenarios. (Bone and joint infections = infections of bones, joints, and related organs; BSI = bloodstream infections; cardiac infections = endocarditis and other cardiac infections; CNS infections = meningitis and other bacterial CNS infections; intra-abdominal infections = peritoneal and intra-abdominal infections; LRI and thorax infections = lower respiratory infections and all related infections in the thorax; bacterial skin infections = bacterial infections of the skin and subcutaneous systems; typhoid, paratyphoid, and iNTS = typhoid fever, paratyphoid fever, and invasive non-typhoidal *Salmonella* spp; UTI = urinary tract infections and pyelonephritis)

For pathogens with hypothetical vaccine profiles (developed by experts or provided in preferred product characteristics), we estimated that a vaccine against *Mycobacterium tuberculosis* that meets WHO’s PPC criteria of 80% efficacy, given to infants, with life-long immunity or boosting, would have averted 0.12 (0.11 - 0.13) million deaths and 4.5 (4.1 - 5.0) million DALYs associated with AMR. An improved vaccine against *Streptococcus pneumoniae* (70% efficacy against bloodstream infections (BSI), meningitis and other bacterial central nervous system infections (CNS), 50% efficacy against lower respiratory infection and all related infections in the thorax (LRI), given to 90% of infants at six weeks of life) would have a relatively highest impact by averting 99 (86 - 115) thousand deaths and 8.6 (7.5 - 10) million DALYs associated with AMR in 2019. An M72-like vaccine against *Mycobacterium tuberculosis* given to adolescents and older populations with life-long immunity or boosting would avert 71 (64 - 78) thousand deaths and 2.6 (2.3 - 2.8) million DALYs associated with AMR. A vaccine against all disease presentations of *Klebsiella pneumoniae* infection given to infants and elderly populations would avert 64 (59 - 72) thousand deaths and 3.7 (3.3 - 4.1) million DALYs associated with AMR.

In the high-potential scenario (see Table 2b), we estimated that vaccination of at-risk individuals across all age groups against *E. coli* - non-diarrhogenic could avert 0.39 (0.37 - 0.40) million deaths and 13 (12 - 13) million DALYs associated with AMR in 2019. Vaccination of at-risk individuals against *Klebsiella pneumoniae* could avert 0.32 (0.31 - 0.34) million deaths and 14 (13 - 15) million DALYs associated with AMR, and vaccination against *Staphylococcus aureus* could avert 0.32 (0.31 - 0.33) million deaths and 11 (10 - 11) million DALYs associated with AMR.

### Vaccine impact on AMR burden by infectious syndrome

Figure 1b shows the vaccine avertable deaths and DALYs attributable to and associated with bacterial AMR for the different infectious syndromes in 2019 at the global level in the baseline scenario. We estimated vaccine avertable mortality associated with bacterial AMR to be highest for lower respiratory infections at 0.16 (0.14 - 0.17) million deaths and 11 (9.6 - 11) million DALYs for the baseline scenario, followed by tuberculosis at 0.12 (0.11 - 0.13) million deaths and 4.5 (4.1 - 5.0) million DALYs and bloodstream infections at 0.11 (0.10 - 0.12) million deaths and 5.6 (5.1 - 6.3) million DALYs in 2019. In the high-potential scenario, vaccine avertable deaths and DALYs were highest for lower respiratory infections, bloodstream infections, and intra-abdominal infections.

For each infectious syndrome, we stratified the vaccine avertable AMR burden for deaths and DALYs by pathogen in the baseline scenario, as shown in Figure 2 (and Figure A1 in the appendix). *Streptococcus pneumoniae, Staphylococcus aureus*, and *Klebsiella pneumoniae* account for most of the vaccine avertable AMR burden associated with lower respiratory infections. *Klebsiella pneumoniae, Acinetobacter baumannii*, and *Escherichia coli* account for most of the vaccine avertable AMR burden associated with bloodstream infections.

**Figure 2.**
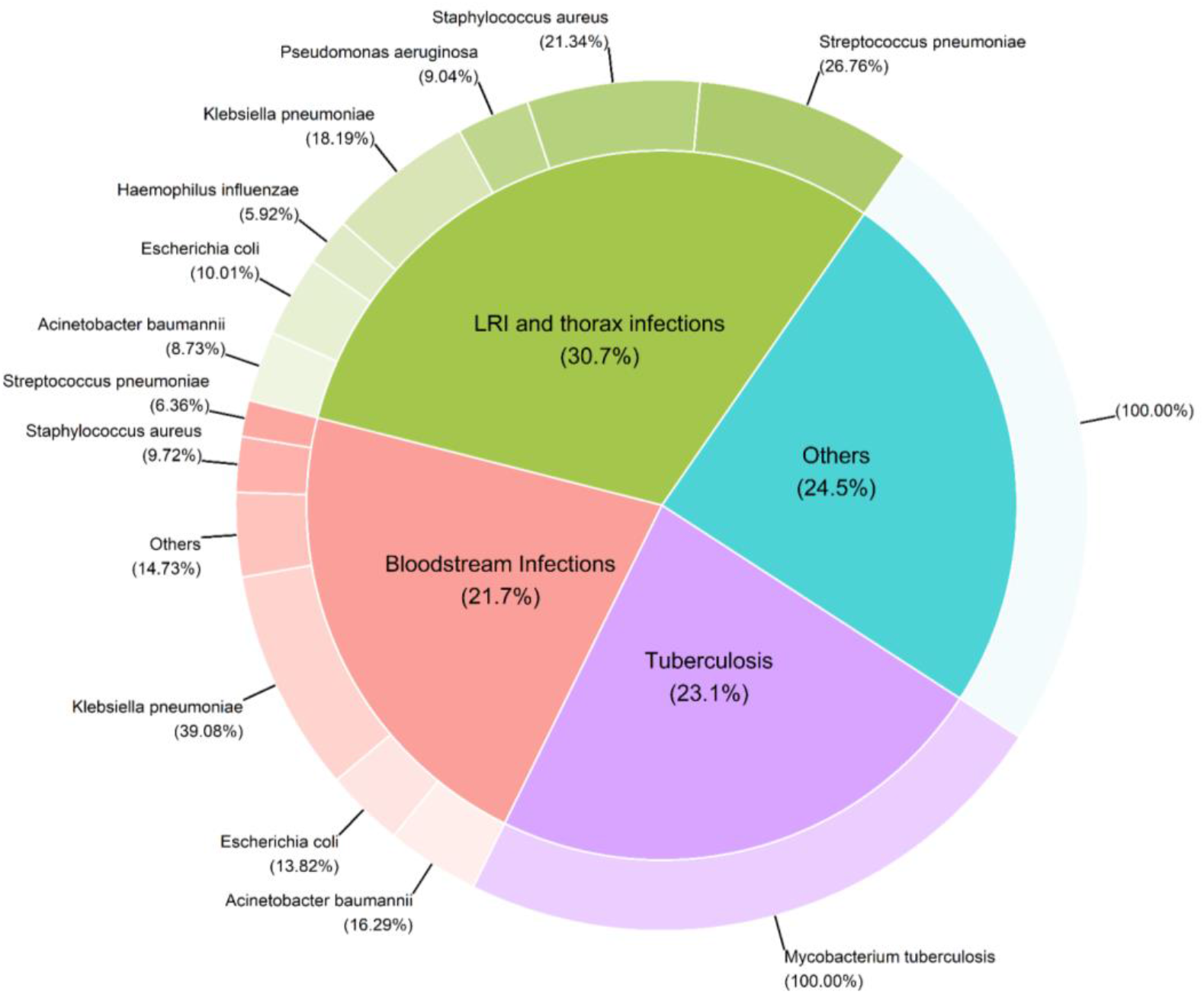
Vaccine avertable AMR burden by infectious syndrome and pathogen. Vaccine avertable deaths associated with AMR by infectious syndrome and pathogen in the baseline scenario. (“Others” include infections of bones, joints, and related organs, bloodstream infections, endocarditis and other cardiac infections, meningitis and other bacterial CNS infections, peritoneal and intra-abdominal infections, lower respiratory infections and all related infections in the thorax, bacterial infections of the skin and subcutaneous systems, typhoid fever, paratyphoid fever, and invasive non-typhoidal Salmonella spp, and urinary tract infections and pyelonephritis)

### Vaccine impact on AMR burden at the regional level

Table 3 and Figure 1c show the vaccine avertable deaths and DALYs attributable to and associated with bacterial AMR at the regional levels in 2019 for the baseline scenario. We estimated the vaccine avertable burden associated with bacterial AMR to be highest in the WHO Africa region at 0.17 (0.15 - 0.18) million deaths and 12 (11 - 13) million DALYs, followed by the WHO South-East Asia region at 0.16 (0.15 - 0.18) million deaths and 7.5 (6.8 - 8.5) million DALYs in 2019. The vaccine avertable AMR burden for the WHO Africa and South-East Asia regions accounts for around two-thirds of the vaccine avertable AMR burden globally in 2019. In the high-potential scenario, we estimated that vaccines would avert an additional 0.19 (0.18 - 0.20) million deaths and 9.6 (8.8 - 11) million DALYs associated with AMR in the WHO Africa region, and 0.32 (0.30 - 0.33) million deaths and 11 (10 - 11) million DALYs associated with AMR in the WHO South-East Asia region.

**Table 3.**
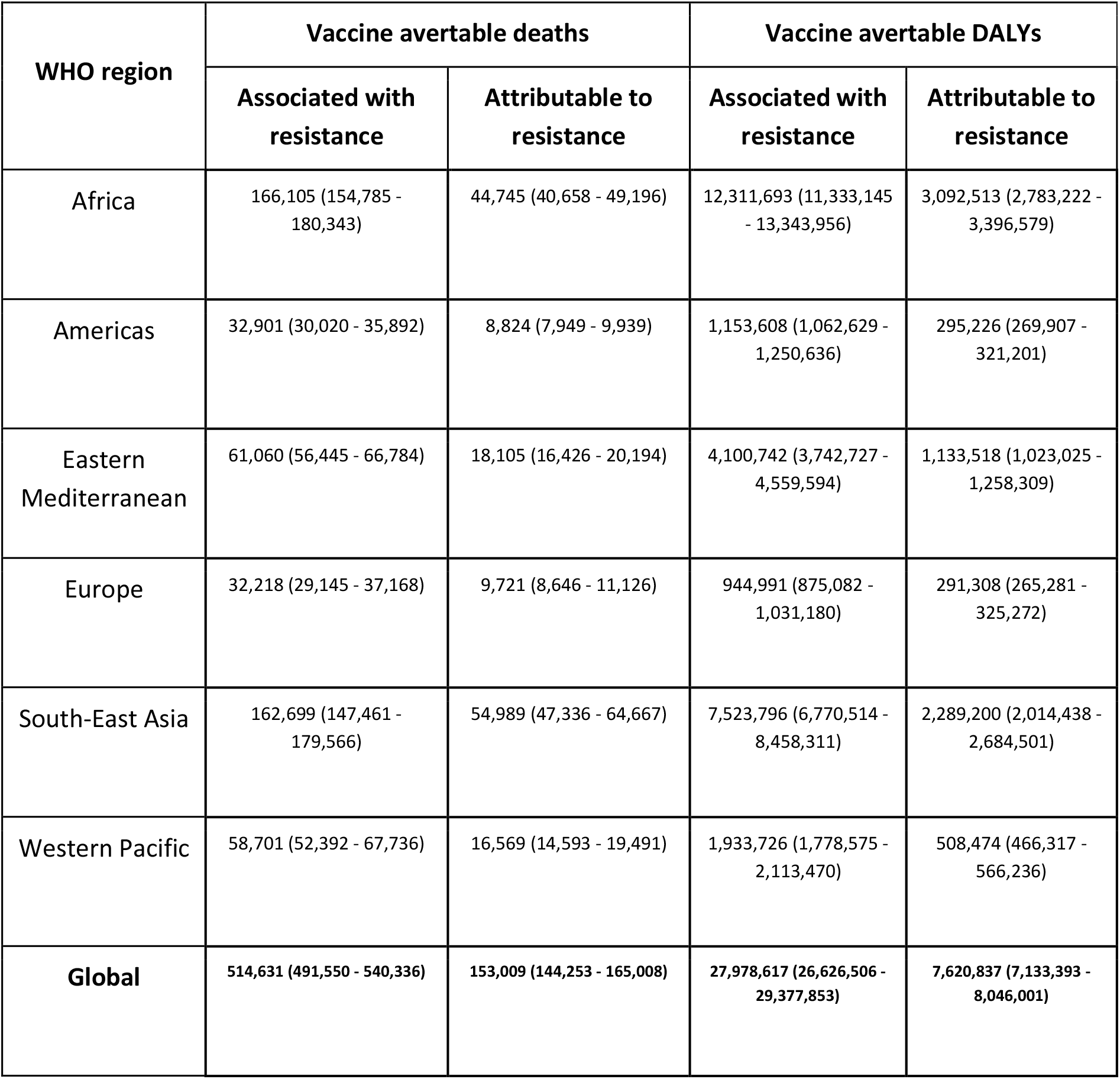
Vaccine avertable AMR burden globally and by WHO region. The estimates (median and 95% uncertainty intervals) for vaccine avertable disease burden attributable to and associated with bacterial AMR in 2019 is presented in terms of deaths and DALYs avertable by vaccination in the baseline scenario.

## Discussion

We estimated vaccine avertable disease burden attributable to and associated with AMR for existing and new vaccines in the pipeline by pathogen, infectious syndrome, and region based on the most recent, comprehensive estimates of the global burden of AMR. The AMR burden avertable by vaccination in 2019 was highest for the WHO Africa and South-East Asia regions, for lower respiratory infections, tuberculosis, and bloodstream infections by infectious syndromes, and for *Mycobacterium tuberculosis* and *Streptococcus pneumoniae* by pathogen.

Our estimates show the impact of existing vaccines for pneumococcal conjugate vaccine (PCV), *Haemophilus influenzae* type b (Hib), and typhoid conjugate vaccine (TCV) on reducing AMR burden attributable to and associated with *Streptococcus pneumoniae, H. influenzae*, and *Salmonella* Typhi respectively. We highlight the critical need to scale up existing vaccines to high and equitable immunisation coverage, and the acceleration of TCV introductions in high burden countries. Also, we show that vaccines can contribute towards preventing a significant proportion of the AMR burden for pathogens which have vaccines in late-stage clinical development with clear attributes or published preferred product characteristics or target product profiles, such as for ExPEC and *Mycobacterium tuberculosis*. Novel regulatory and policy mechanisms should be developed to accelerate the approval and use of these vaccines to prevent AMR. Based on the estimated high vaccine avertable burden associated with AMR for *Klebsiella pneumoniae, Staphylococcus aureus*, and *Acinetobacter baumannii*, we urgently call for studies to enhance biological understanding and improve the feasibility of developing vaccines for these pathogens. For the remaining pathogens that have vaccine candidates in the early stages of clinical development or no vaccines in the pipeline, we recommend investing in vaccine development to resolve biological challenges as well as feasibility in terms of product development, market access, and product implementation.

Our analysis included a baseline and high-potential scenarios. In the baseline scenario, we model vaccine delivery based on known vaccine attributes, including a defined target age group that has been immunised with a vaccine in the past, during clinical trials, or identified in vaccine target product profiles. In contrast, the high-potential scenario makes no assumptions about vaccine delivery and target age group and shows the highest probable vaccine impact, should there be a policy recommendation and feasibility of delivery to all who would benefit from a vaccine. We recognise that the high-potential scenario includes multiple challenges that need overcoming such as immunisation of adults and the elderly, timely immunisation to prevent nosocomial infections, vaccine efficacy in patients who are immunocompromised and with co-morbidities, vaccine demand, and financing.

Pan-pathogen analyses with standardised methodologies are critical to inform vaccine funding and development and should be followed up with detailed vaccine-specific analyses, considering pathogen biology and transmission, and accounting for varied disease burden patterns across the spatial and temporal scales. *Haemophilus influenzae* type b (Hib), rotavirus, pneumococcal, typhoid and influenza vaccines have been directly associated with reduction of resistance, antibiotic use and related clinical complications [9,15–22], while Fu *et al*. modelled the global burden of drug-resistant tuberculosis avertable by a future TB vaccine [23].

Our study has limitations. First, since we included the direct effect of vaccination but excluded indirect effect and transmission dynamics of AMR pathogens, our vaccine impact estimates on averted AMR burden are conservative. Second, our analysis focused on 15 bacterial pathogens and additional pathogens included in the GRAM project such as *Enterobacter* spp., Group B *Streptococcus, Enterococcus feacalis, Proteus* spp., *Citrobacter* spp., and *Morganella* spp. were excluded. However, inclusion of these pathogens appears unlikely to significantly affect our overall inferences considering that the included 15 pathogens are responsible for the majority of the AMR burden. Third, while our analysis was based on the estimates generated by the GRAM project, which represents the most comprehensive estimates of bacterial AMR burden to date, limited input data to the GRAM project especially from low- and middle-income countries is a significant data gap that necessitates newer surveillance data and platforms to inform the updates, validity, and confidence in the estimates of the GRAM project. In particular, estimates from the GRAM project for tuberculosis do not include tuberculosis associated with HIV. Fourth, we did not consider the impact of viral vaccines on reducing the AMR drivers of antibiotic misuse and overuse [18,20,24,25]. Finally, we did not consider geographic and socioeconomic clustering of vaccination coverage, which could lead to heterogeneity in vaccination impact on lowering AMR burden with relatively less impact among subpopulations with higher risk of disease while also facing lower health care access including access to vaccination services [26].

The value of vaccines in preventing AMR should be systematically considered in the decision-making process during scale-up of existing vaccines and introduction of new vaccines. Vaccines should be explicitly incorporated as tools to combat AMR into National Action Plans on AMR [27] and National Immunisation Strategies [28]. For new vaccines in the pipeline and future vaccines, we recommend vaccine avertable burden of AMR to be included in the full value of vaccine assessments [29]. This evidence can support stakeholders in their decision-making process and priority setting throughout the end-to-end continuum from discovery and clinical development to investment, development, introduction, and sustainability of new vaccines with equitable access.

## Data Availability

All data produced in the present study are available upon reasonable request to the authors.

## Ethics approval

This study was approved by the ethics committee (Ref 26896) of the London School of Hygiene & Tropical Medicine.

## Data availability and code repository

We conducted our analysis using the R programming language for statistical computing [30], and the repository for the data and software code of this modelling study are publicly accessible at https://github.com/vaccine-impact/vaccine_amr.

## Contributors

CK, MH, IF, MHA, and KA conceptualised and designed the study. IF and MHA compiled the data sets. CK developed the proportional impact model and software and conducted the analysis. CK and KA wrote the first draft, and all authors contributed to reviewing and editing of the manuscript and have approved the final version.

## Acknowledgements

We thank Mohsen Naghavi, Gisela Robles Aguilar, and Eve Wool of the Institute for Health Metrics and Evaluation for sharing the bacterial AMR burden data. We thank Ramanan Laxminarayan (Center for Disease Dynamics, Economics & Policy) and Padmini Srikantiah (Bill & Melinda Gates Foundation) for helpful discussions. We thank John Clemens, Nimesh Poudyal, and Raphael Zellweger at the International Vaccine Institute for their review and feedback on this article. We appreciate the feedback on this article from the members of the WHO Technical Advisory Group on Vaccines and Antimicrobial Resistance (TAG VAC-AMR), including Holy Akwar (International Vaccine Institute), Kate Baker (University of Liverpool), Sulagna Basu (Indian Council of Medical Research - National Institute of Cholera and Enteric Diseases), Julie Bines (University of Melbourne), Jeremy Brown (University College London), Bill Hausdorff (PATH), Karen Keddy, Kirsty Ledoare (Imperial College London), Shelley Magill (Centers for Disease Control and Prevention), Mohsen Naghavi (Institute for Health Metrics and Evaluation), Olga Perovic (National Institute for Communicable Diseases and University of the Witwatersrand), Manish Sadarangani (University of British Columbia), and Johan Vekemans (International AIDS Vaccine Initiative). We thank Kevin Ikuta (University of California Los Angeles and University of Washington) and Tomislav Meštrović (Institute for Health Metrics and Evaluation) for their feedback on this article.

## Patient and Public Involvement

Patients or the public were not involved in the design, or conduct, or reporting, or dissemination plans of our research.

### Appendix

#### A1. Additional details of the modelling process

##### Estimating vaccine-avertable AMR burden of the target age group

The AMR burden data from the Global Research on Antimicrobial Resistance (GRAM) project was disaggregated by age and included the following categories – early neonatal (first week after birth), late neonatal (2-4 weeks of age), postneonatal (5 weeks to under 1-year of age), 1-4 years, 4-9 years, …, 90-94 years, and 95 years and beyond. We estimated the reduction in AMR burden in direct proportion to efficacy, coverage, target population for protection, and duration of protection of existing and potential future vaccines. We considered that immunised individuals would gain vaccine-derived immunity two weeks post-vaccination.

##### Estimating pre-vaccination burden for pathogens with existing vaccines

For the existing Hib and pneumococcal conjugate vaccines (PCVs), we estimated the pre-vaccination (i.e., no vaccination) burden associated and attributable to AMR in 2019 using the estimates of efficacy and coverage in 2019. We used the vaccine coverage for Hib and PCV from WHO/UNICEF Estimates of National Immunization Coverage (WUENIC) [1] and demography data from the United Nations World Population Prospects (UNWPP) [2] to estimate the vaccine coverage at the regional level. We used the vaccine efficacy estimates for the first dose, second dose, and third doses scheduled at 6, 10, and 14 weeks for the Hib [3,4] and PCV vaccines [5,6]. By applying the vaccine efficacies and regional coverage to the AMR burden data in 2019, we estimated the increase in AMR burden for the counterfactual scenario of no vaccination in direct proportion to efficacy, coverage, target population for protection, and duration of protection. The global and regional coverage of typhoid conjugate vaccine and the post-vaccination impact was minimal in 2019 [7] and thereby did not warrant additional estimation for the counterfactual scenario of no vaccination.

The GRAM project estimates of AMR burden for *H. influenzae* were not stratified by serotypes. *H. influenzae* serotype b (Hib) was responsible for around 95% of all invasive *H. influenzae* disease burden among children younger than 5 years of age before the introduction of vaccines [8]. By applying the 95% Hib proportion to the total *H. influenzae* burden in the counterfactual scenario of no vaccination, we estimated the vaccine-preventable proportion of Hib-specific AMR burden of the total *H. influenzae* AMR burden in 2019.

##### Disease type specification of the AMR burden

The GRAM project’s AMR burden estimations do not differentiate between *Escherichia coli* strains. Instead, the AMR burden estimates were stratified by symptoms. Since enterotoxigenic *E. coli* (ETEC) and enteropathogenic *E. coli* (EPEC) are the two major *E. coli* strains that cause diarrhoea, we calculated the proportional contribution of ETEC to the AMR burden due to *E. coli* causing diarrhoea and then estimated the impact of the ETEC vaccine on reducing this burden (43.97%).

##### Estimating the aggregated vaccine avertable burden

To estimate the aggregate estimates on the impact of the vaccines by region and by infectious syndrome, we estimated the impact of all listed vaccines as long as the effects do not overlap to avoid double counting. When there were multiple vaccines which target the same disease, infectious syndrome, and age, we chose the vaccines with greater efficacy for these estimates. However, for vaccines against *Streptococcus pneumoniae*, we used the efficacy of the existing vaccine with increased coverage that met the strategic priority on coverage and equity of Immunisation Agenda 2030.

##### Scenarios for vaccine avertable AMR burden

We estimated vaccine avertable AMR burden for baseline and high-potential scenarios. We recognise that the high-potential scenario is optimistic given the unanswered questions about the feasibility of producing vaccines with long-term immunity and timely delivery to populations at risk, such as patients in hospitals undergoing elective surgeries. We included the high-potential scenario to highlight the potential impact vaccines could have if challenges around vaccine development and delivery were to be resolved.

##### Uncertainty analysis

Our estimations account for the uncertainties around AMR burden, efficacy, and coverage. Based on data examination, we applied the lognormal distribution to the mean, the 2.5th and 97.5th percentiles of the AMR burden to generate the randomly drawn values. For vaccine efficacy and coverage, we used the truncated normal distribution. For hypothetical vaccines, we applied ± 20% to the vaccine efficacy and coverage on the vaccine profile. For existing vaccines, we used confidence intervals of the vaccine efficacy from studies and applied ± 5% to vaccine coverage (that is, coverage of existing vaccines increased in order to meet the strategic priority on coverage and equity of Immunisation Agenda 2030). When estimating the impact of the existing vaccines with current coverage (that is, based on WUENIC estimates), we only included the uncertainty in efficacy as we used the point estimates of actual coverage.

##### Data availability and code repository

We conducted our analysis using the R programming language for statistical computing [28], and the repository for the data and software code of this modelling study are publicly accessible at https://github.com/vaccine-impact/vaccine_amr.

**Figure A1.**
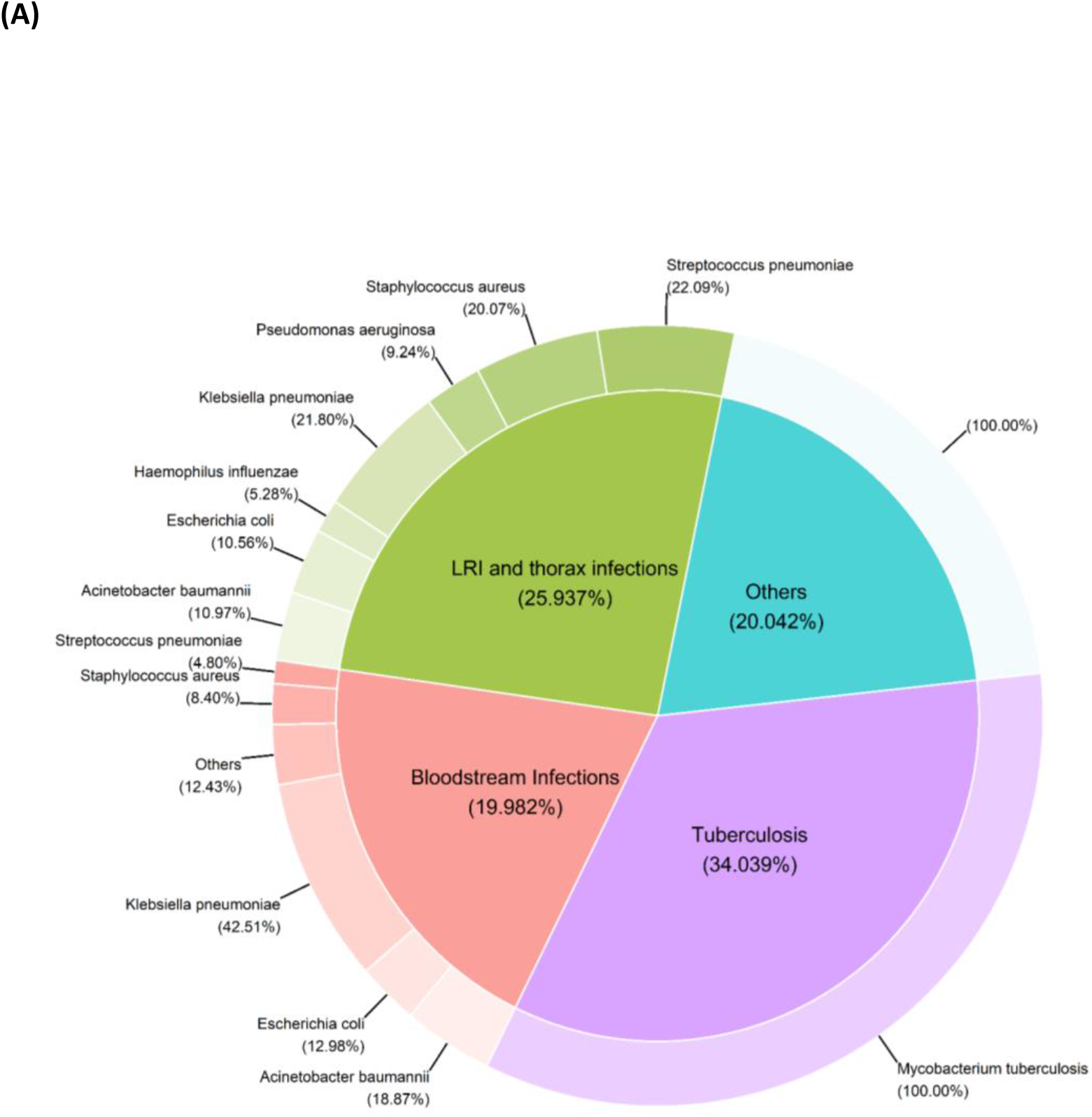
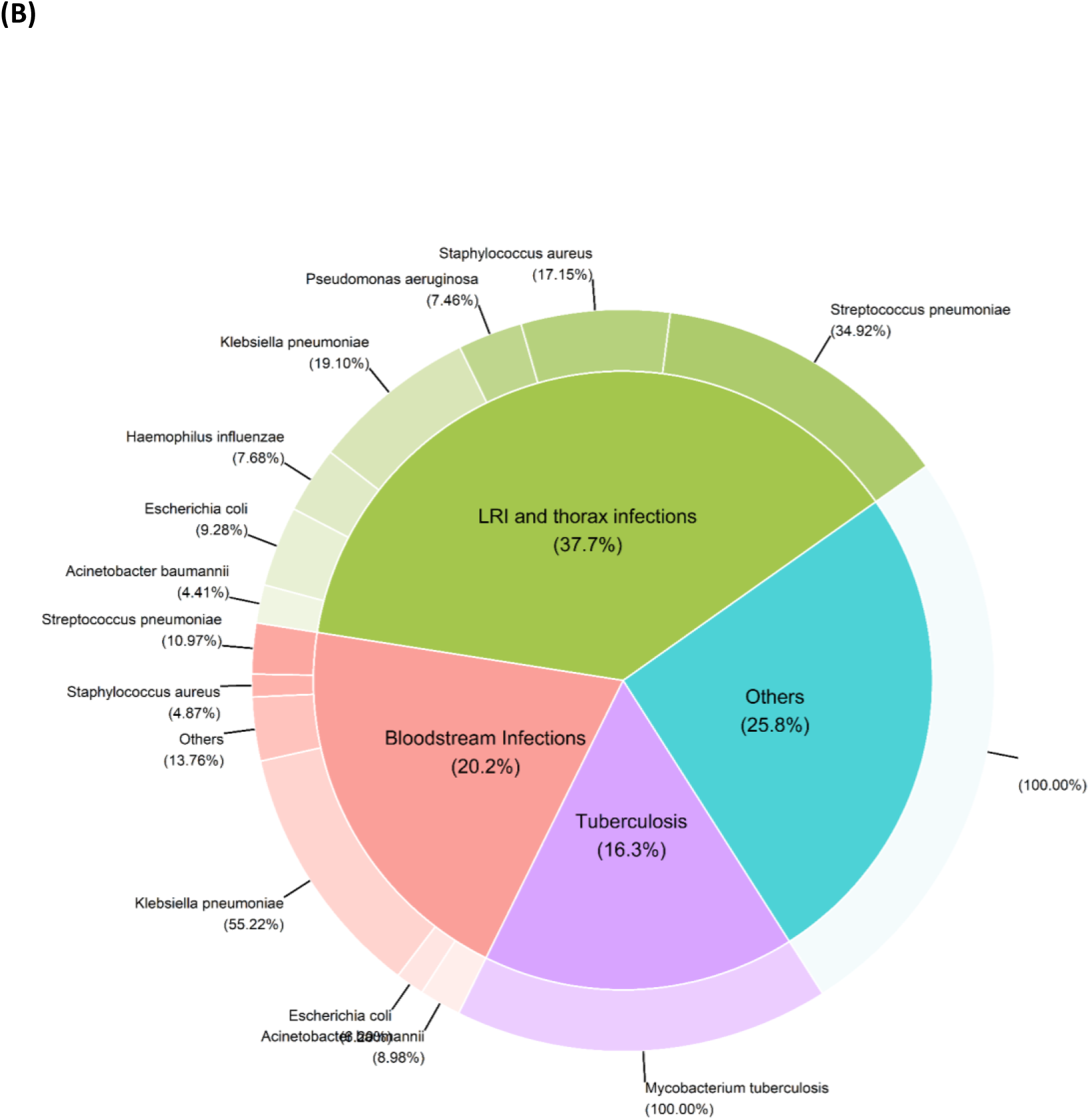
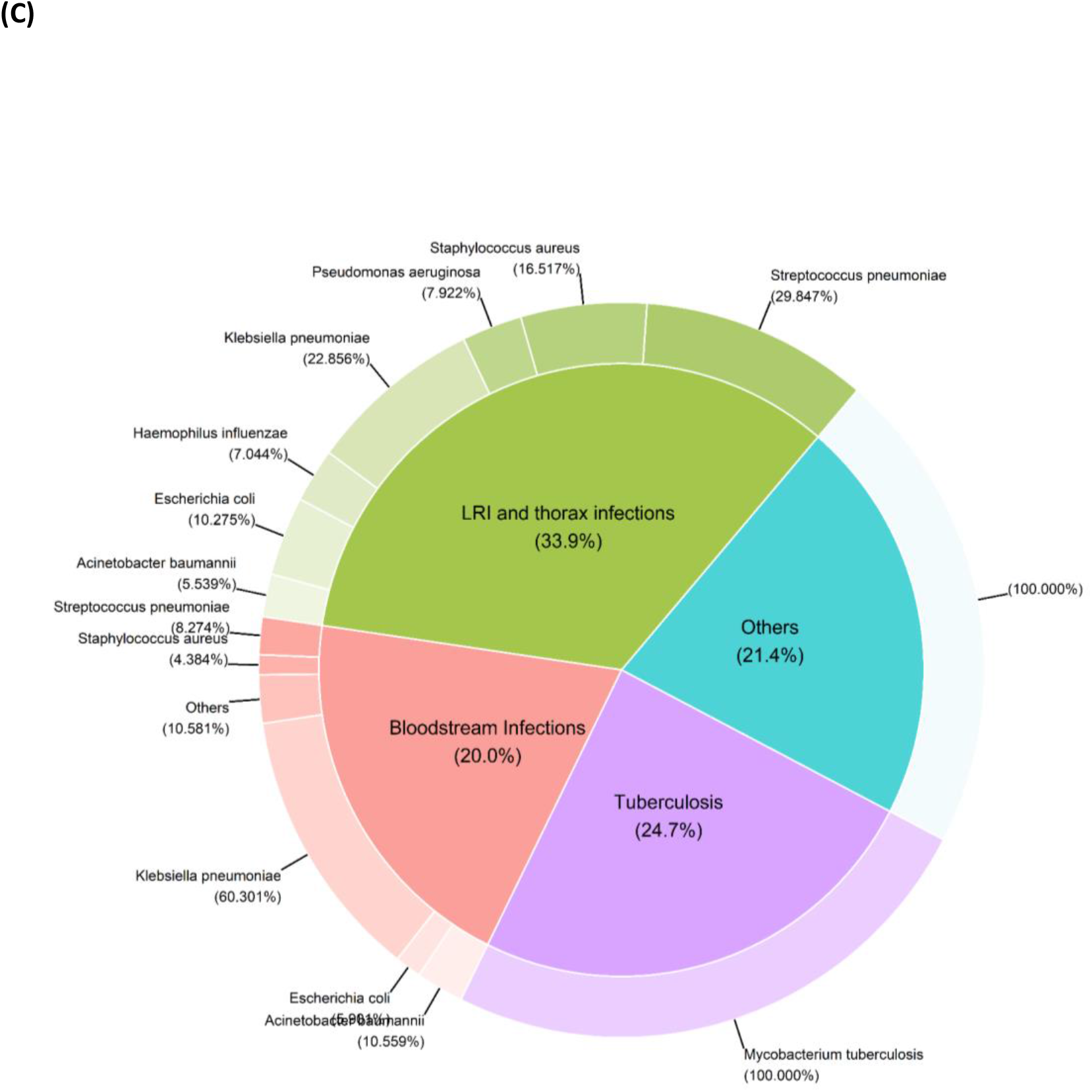
Vaccine avertable AMR burden by infectious syndrome and pathogen. Vaccine avertable AMR burden (deaths and DALYs averted) by infectious syndrome and pathogen in the baseline scenario. (A) Vaccine avertable deaths attributable to AMR by infectious syndrome and pathogen. (B) Vaccine avertable DALYs associated with AMR by infectious syndrome and pathogen. (C) Vaccine avertable DALYs attributable to AMR by infectious syndrome and pathogen.

